# Evaluation of serum antibodies against SARS-CoV□2 in healthcare workers who participated in the operation of charter flights for the evacuation of Japanese residents from Hubei Province

**DOI:** 10.1101/2021.03.17.21251964

**Authors:** Tetsuya Suzuki, Kayoko Hayakawa, Akira Ainai, Naoko Iwata-Yoshikawa, Kaori Sano, Noriyo Nagata, Tadaki Suzuki, Yuji Wakimoto, Yutaro Akiyama, Yusuke Miyazato, Keiji Nakamura, Satoshi Ide, Hidetoshi Nomoto, Takato Nakamoto, Masayuki Ota, Yuki Moriyama, Yuko Sugiki, Sho Saito, Shinichiro Morioka, Masahiro Ishikane, Noriko Kinoshita, Satoshi Kutsuna, Norio Ohmagari

## Abstract

There are several recommendations for the use of personal protective equipment (PPE) against severe acute respiratory syndrome coronavirus 2 (SARS-CoV-2) infection. However, the selection of appropriate PPE for the current situation remains controversial. We measured serum antibody titers for SARS-CoV-2 in 10 participants who were engaged in the operation of charter flights for the evacuation of Japanese residents from Hubei Province. All participants wore PPE in accordance with Centers for Disease Control and Prevention guidelines. A total of 17 samples were tested, and all were seronegative. Hence, we conclude that the current PPE recommendation is effective to protect healthcare workers from SARS-CoV-2 infection.

## Text

Although there are several recommendations for personal protective equipment (PPE) against the severe acute respiratory syndrome coronavirus 2 (SARS-CoV-2) infection (1, 2), the selection of appropriate PPE according to the current situation remains a matter of debate. There are reports on the incidence of coronavirus disease (COVID-19) among healthcare workers (3), which puts them under tremendous stress. The National Center for Global Health and Medicine (NCGM) helped the operation of charter flights for the evacuation of Japanese residents from Hubei Province from the end of January to mid-February 2020 (4). In the present study, we evaluated the efficacy of PPE against SARS-CoV-2 infection among hospital staff who were engaged in the operation of charter flights by measuring their serum antibody levels. In addition, we used a questionnaire for self-evaluation of the PPE wearing status.

Hospital staff who participated in the operation were recruited for this study. All staff wore PPE, which includes N95 mask, gown, gloves, disposable cap, and eye protection, in accordance with the Centers for Disease Control and Prevention (CDC) guidelines. The details of this operation are provided in a previous report (4). The doctor who collected the pharyngeal swab had used a coverall to minimize the risk of body surface contamination due to infected secretions and droplets. Enhanced hand hygiene using an alcohol-based hand sanitizer was practiced.

Blood samples were collected at enrollment (after February 14th) and at every 2 weeks after enrollment until 4 weeks after the final participation in the evacuation operation. At enrollment, participants were asked to answer the web-form self-reporting questionnaire about their PPE and their compliance. This study was reviewed and approved by the Ethics Committee of the Center Hospital of National Center for Global Health and Medicine. Written informed consent was obtained from all participants.

Serum antibody titers were measured using a two-step assay. First, samples were screened using enzyme-linked immunosorbent assay (ELISA). Positive samples were then re-examined, by neutralization assay, for the presence or absence of specific antibodies against SARS-CoV-2. The details of this method have been reported previously (5). The participants of the previous study and this present study are not overlapped.

Japanese government-chartered aircrafts were operated on 3 times in January and twice in February, 2020. The number of people who were screened at our hospital on the respective dates was 199, 197, 140, 194, and 63, respectively. The number of positive cases was 3 (1), 2 (3), 2 (1), 1, and 0, respectively (numbers in parentheses indicate the number of people whose results were initially negative from the samples obtained at NCGM but later became positive) (6-9). Thus, the final chance for participants to come in contact with COVID-19 patients was in early February, but two participants had other chances of occupational exposure to COVID-19 in February.

Ten healthcare workers (2 nurses, 5 doctors, and 3 clerical staff) were enrolled. Five of them were male. Eight participants responded to a web-form questionnaire. Their characteristics are listed in Table 1. Among those who answered the questionnaire, 4 were in their 40s, 3 were in their 30s, and 1 was in his/her 50s. Their median compliance with PPE was 90% (range 70-100%, n=8). A total of 17 blood samples were collected from 10 participants. Of them, 8 had their first samples collected around late February, 2020. Before blood sampling, 25% of them (n=2/8) noticed some respiratory symptoms. Six participants underwent more than 2 blood samplings at intervals of approximately 2 weeks. Five samples from 2 participants tested positive on ELISA screening, but none of them remained positive after the neutralization assay. Thus, all samples from all participants were seronegative.

**Table 1.**
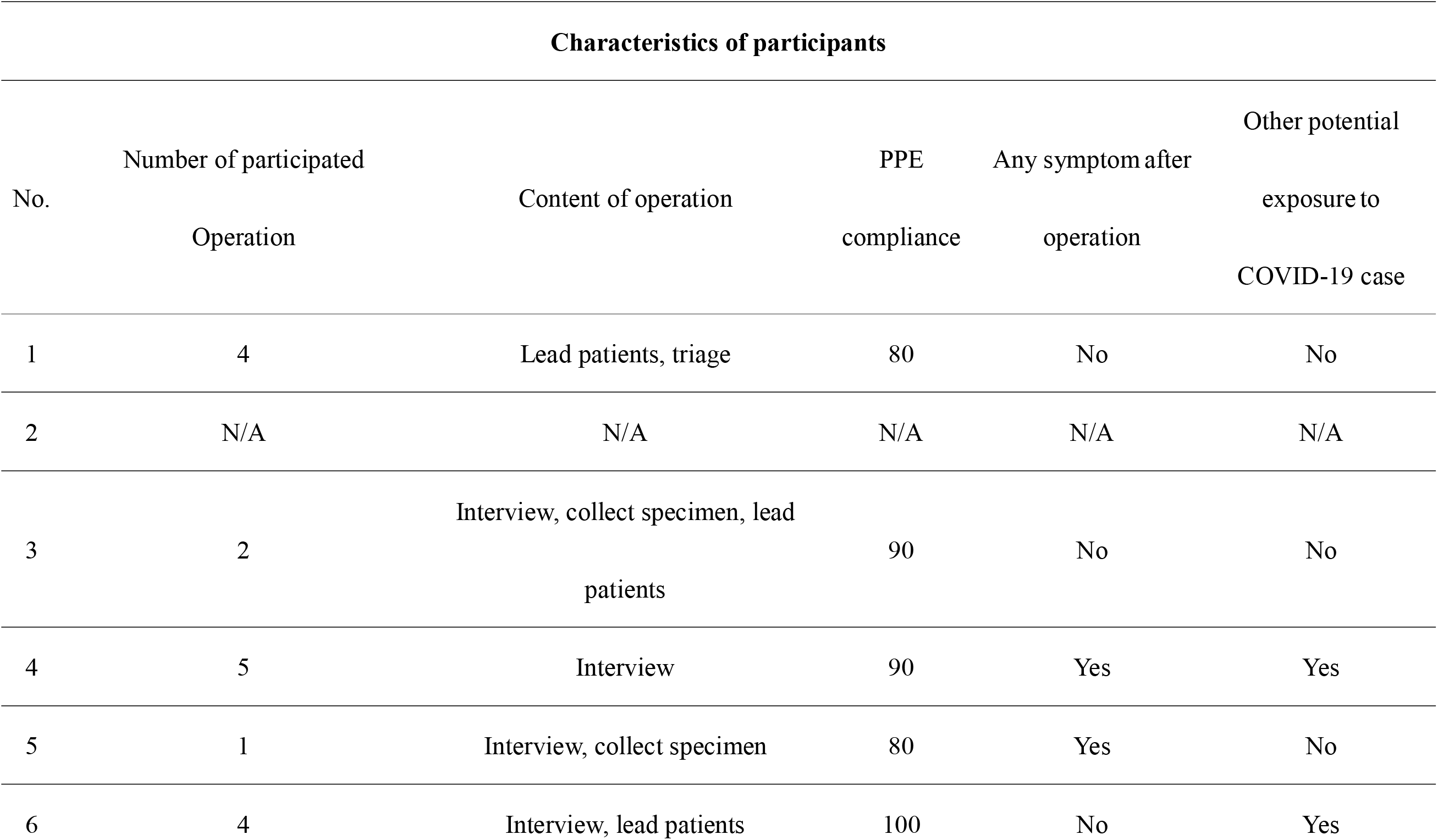
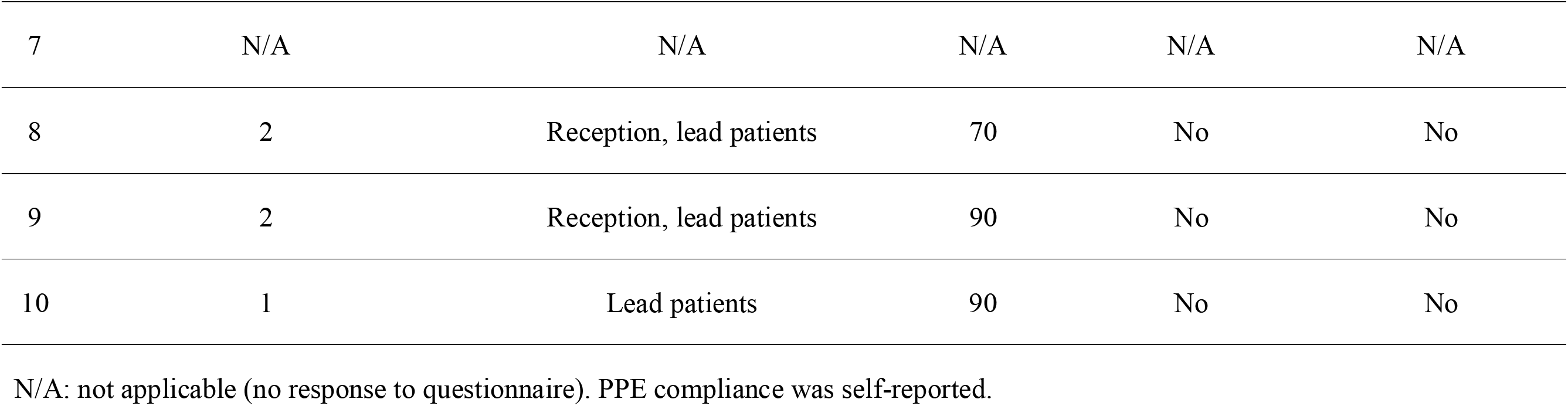
Characteristics of participants and timing of blood sampling.

| Characteristics of participants |  |  |  |  |  |
| --- | --- | --- | --- | --- | --- |
| No. | Number of participated<br>Operation | Content of operation | PPE<br>compliance | Any symptom after<br>operation | Other potential<br>exposure to<br>COVID-19 case |
| 1 | 4 | Lead patients, triage | 80 | No | No |
| 2 | N/A | N/A | N/A | N/A | N/A |
| 3 | 2 | Interview, collect specimen, lead<br>patients | 90 | No | No |
| 4 | 5 | Interview | 90 | Yes | Yes |
| 5 | 1 | Interview, collect specimen | 80 | Yes | No |
| 6 | 4 | Interview, lead patients | 100 | No | Yes |
| 7 | N/A | N/A | N/A | N/A | N/A |
| 8 | 2 | Reception, lead patients | 70 | No | No |
| 9 | 2 | Reception, lead patients | 90 | No | No |
| 10 | 1 | Lead patients | 90 | No | No |

N/A: not applicable (no response to questionnaire). PPE compliance was self-reported.

We conclude that healthcare workers who participated in the operation of charter flights for the evacuation of Japanese residents from Hubei Province, China, were seronegative for SARS-CoV-2. Thus, PPE in accordance with the CDC guidelines is effective in protecting healthcare staff from COVID-19. Along with previous studies (10, 11), these results suggest that appropriate PPE prevents healthcare workers from contracting COVID-19. The unique aspects of our study are as follows: (a) participants included clerical staff who are usually not familiar with the use of PPE and (b) a short period of training would enable them to use PPE appropriately. In addition, to maximize the sensitivity and specificity, we used a careful, two-step evaluation for the serostatus.

In our study, the first blood sample was obtained between late February and early March, 2020. Because there were no laboratory-confirmed COVID-19 patients in the 5^th^ flight, their final potential exposure was on early February, 2020. The duration was sufficient to attain a seropositive status, if the infection was established among the participants (12).

There are some limitations to this study. First, recall bias is inevitable because the questionnaire used was self-reporting. Second, compared to inpatient care, opportunities for exposure to COVID-19 cases were less intense in this cohort. Third, the setting of this study is different from the routine medical exposure, and thus, the results of our study might not be adaptable to clinical practice.

In conclusion, this study demonstrates the effectiveness of PPE in protecting a wide range of healthcare workers from being infected with SARS-CoV-2. Sufficient PPE supply for healthcare staff is essential to avoid the spread of nosocomial infections. Further studies including a large number of participants are required to support and validate the effectiveness of PPE.

## Data Availability

The datasets of the current study are available from the corresponding author on reasonable request.

## Ethical Approval

This study was reviewed and approved by the Ethics Committee of the Center Hospital of the National Center for Global Health and Medicine. Written informed consent was obtained from all participants.

## Acknowledgement

The authors acknowledge BZ, KT, FN, NM, JL, YG, YO, JK, ST, and BS for their help in sample collection.

